# Towards the global equilibrium of COVID-19: statistical analysis of country–level data

**DOI:** 10.1101/2021.08.23.21262413

**Authors:** Mark Last

## Abstract

**Objectives:** The time-varying effect of COVID-19 on a population of a given country or territory can be measured by the Reproduction Number (R) and the Case Fatality Rate (CFR). In our study, we explore the dynamics of these two measures to test whether the virus has reached its equilibrium point and to identify the main factors explaining the current R and CFR variability across countries.

**Design:** A retrospective study of publicly available country-level data.

**Setting:** Fifty countries having the highest number of confirmed COVID–19 cases at the end of September 2021.

**Participants:** Aggregated data including 213 976 306 COVID-19 cases confirmed in the selected fifty countries from the start of the epidemic to September 30, 2021.

**Primary and secondary outcome measures:** The daily values of COVID-19 R and CFR measures were estimated using country-level data from the Our World in Data website.

**Results:** The mean values of country–level moving averages of R and CFR went down from 1.118 and 6.3%, respectively, on June 30, 2020 to 1.083 and 3.6% on September 30, 2020 and to 1.015 and 1.8% by September 30, 2021. In parallel, the 10% to 90% inter-percentile range of R and CFR moving averages decreased from 0.288 and 13.3%, respectively, on June 30, 2020, to 0.151 and 7.7% on September 30, 2020, and to 0.107 and 3.3% by September 30, 2021. According to a comparison of the country–level 180–day moving averages of R and CFR calculated on September 30, 2021, an increase of 1% in the Delta variant share is associated with an increase of 0.0009 (95% CI 0.000 to 0.002) in the average Reproduction Number R, while an increase of 1% in the total percentage of confirmed COVID-19 cases per country’s population is associated with a decrease of 0.005 (95% CI 0.000 to 0.010) in the average R. Also, an increase of 1% in the total percentage of fully vaccinated people per country’s population is associated with a decrease of 0.04% (95% CI 0.01% to 0.06%) in the average CFR. Other virological, demographic, economic, immunization, or stringency factors were not statistically significantly associated with either R or CFR across the explored countries.

**Conclusions:** The slow decrease in the country-level moving averages of R, approaching the level of 1.0 and accompanied by repeated outbreaks (“waves”) in various countries, may indicate that COVID-19 has reached its point of a stable endemic equilibrium. A regression analysis implies that only a prohibitively high level of herd immunity (about 63%) may stop the endemic by reaching a stable disease-free equilibrium. It also appears that fully vaccinating about 70% of a country’s population should be sufficient for bringing the CFR close to the level of a seasonal flu (about 0.1%). Thus, while the currently available vaccines prove to be effective in reducing the mortality from the existing COVID-19 variants, they are unlikely to stop the spread of the virus in the foreseeable future. It is noteworthy that no statistically significant effects of government measures restricting the people’s behavior (such as lockdowns) were found in the analyzed data.

**Strengths and limitations of this study:**

- In this study, we have explored the long–term trends in country–level Reproduction Number and the Case Fatality Rate of COVID-19.
- Our study also investigated the long–term statistical dependence of the COVID-19 Reproduction Number and the Case Fatality Rate on epidemiological, demographic, economic, immunization, and government policy factors in each country.
- The findings of this study may have important implications for the health policy-makers worldwide.
- The officially reported numbers of daily COVID-19 confirmed cases depend on the local testing policy and usually underestimate the true number of carriers in the population.
- The officially reported numbers of daily COVID-19 deaths in some countries may include all deceased individuals who tested positive for COVID-19, disregarding their actual cause of death, and exclude victims who were not tested for COVID-19.

## Introduction

The first case of COVID-19 was reported in the Chinese city of Wuhan in December 2019. According to the Humanitarian Data Exchange website (HDX 2020), on January 22, 2020 there were 557 confirmed cases of COVID-19 in 29 different countries. On the same date, the global number of reported COVID-19 victims has reached 17, all of them from the Hubei province in China. Due to the extensive international travel, the virus has spread quickly around the world, exceeding one million cases in 254 countries and territories by the end of March 2020 and 100,000 deaths 8 days later. The continuous response of national and regional authorities to the pandemic varied significantly from the near-absolute closure of international borders (e.g., Australia and New Zealand) and repeated lockdowns (e.g., New York City and Israel) to focusing on the protection of high–risk population only (e.g., Sweden). As the pandemic continued to spread, each period of a steady increase in either local or global amount of COVID-19 cases and deaths was always followed by an opposite, decreasing trend, frequently assumed to be a result of various intervention measures. However, in many cases, another, often a deadlier “wave” took place some time later. Starting from December 2020, many countries, in the hope of “winning the pandemic”, have launched massive vaccination campaigns, which continue at the time of writing this article. By the end of July 2021, 1.14 billion people (14.6% of the world population) were fully vaccinated, while the global number of confirmed COVID-19 cases approached 200 million with about 4.2 million victims in 251 different countries and territories.

Considering the widespread travel restrictions at the time of the pandemic, the COVID-19-related death toll in a given country depends mainly on the following two factors: the average value of the effective (time–varying) reproduction number *R*_*e f f*_, or *R*_*t*_, which represents the average number of cases an infected person has generated in the country’s population, and the average Case Fatality Rate (CFR), calculated as a percentage of death outcomes out of all cases confirmed during a specific period. Thus, we focus our study on the comparative analysis of these two parameters at the country level.

In a completely susceptible population, the effective reproduction number *R*_*e f f*_ equals to the basic reproduction number *R*_0_, defined as the average number of secondary infections an infected person will cause in an “immunologically naive” population before he or she is effectively removed from that population as a result of recovery, hospitalization, quarantine, etc. (Maier and Brockmann 2020). The population is expected to reach “herd immunity” when the proportion of non-susceptible (“immunologically experienced”) individuals exceeds 1 − 1/*R*_0_ (Kadkhoda 2021). A direct measurement of *R*_0_ requires identifying the exact source of each infection case, which is rarely possible. However, the basic reproduction number can be estimated from the epidemiological data using a mathematical model such as SEIR (susceptible–exposed–infected–recovered) (Anderson and May 1992). The authors of (Linka et al. 2020) used the SEIR model and the reported COVID-19 cases in each one of the 27 the European Union countries for projecting the effective reproduction number *R*(*t*) and predicting the epidemic evolution from May 10 to June 20, 2020. They evaluated three possible scenarios for their prediction period: a constant value of the effective reproduction number *R*(*t*), a slow return to the basic reproduction number *R*_0_ within three months, and a fast return to *R*_0_ within one month. Their study shows that the severe mobility restrictions on air travel, driving, walking and transit, which were implemented across Europe during March - May 2020, were highly correlated with *R*_0_ in most countries, resulting in a drastic reduction of the population weighted mean of the basic reproduction number from 4.22 (CI 2.53 − 5.91) to 0.67 (CI 0.49 − 0.85). The study did not explore additional factors, which could contribute to the *R*_0_ decrease during the same period, such as public health measures, a reduced amount of social interaction (also known as “social distancing”), and natural immunity acquired by the recovered individuals.

In our previous work (Last 2020), we have explored the overall evolution of the basic reproduction number in Israel, Greece, Italy, and Sweden between March and July 2020 using the relationship between the daily reproduction numbers *R*_*t*_, the basic reproduction number *R*_0_(*t*), and the cumulative percentage of confirmed cases *p*_*t*_, which is shown in Eq. 1.

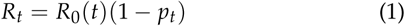

On each day, *R*_*t*_ was estimated under the assumption that the number of deaths on day *t* represents the actual number of new infections on day *t* − 16, which is likely to exceed the number of cases detected through testing. We have shown that some intervention measures taken by the Israeli government, such as mandating mask–wearing in public (April 1) and Passover lockdown (April 7) were actually introduced after the value of *R*_0_(*t*) has already decreased from its starting level of 1.20 to less than the critical level of 1.0, a trend which continued for the succeeding weeks despite a fast relaxation of most restrictions enforced in March and April. After reaching an extremely low level of 0.30 around May 17 and shortly after reopening of all schools in the country (which stayed open until the end of the school year on June 20), the basic reproduction number in the Israeli population started another continuous climb as part of the COVID-19 “second wave”. However, we could not reveal in (Last 2020) a consistent effect of any specific measures on the actual infection rate dynamics.

A systematic review of 108 *R*_0_ estimates reported in 45 articles published between January 1 and August 31, 2020 is presented by (Ahammed *et al*. 2021). The pooled value of *R*_0_ was found to be 2.69 (CI 2.40, 2.98), with statistically insignificant differences between Asian, European, and North-American countries. The review found only a few studies covering Africa and South America. The authors indicate that in the initial stages of a new epidemic, the calculated *R*_0_ values may be overestimated due to insufficient amount of available data. They also explored 158 estimates of CFR presented in 34 articles. The pooled value of the CFR was 2.67% (2.25%, 3.13%), with higher mean CFR values observed for the countries with lower tests (3.15% vs 2.16%) and greater median population age (3.13% vs 2.27%).

The authors of (Cao *et al*. 2020) tried to identify the main factors affecting the case fatality rates in 209 countries and territories based on the COVID-19 data downloaded from the Our World in Data website (Ritchie *et al*. 2020) on 2 July 2020 (including 10,445,656 confirmed COVID-19 cases and 511,030 deaths). They found the average of value of CFR to be about 2%–3% worldwide. The factors directly associated with country-level CFR included the population size and the proportion of female smokers, whereas the open testing policies, cardiovascular disease death rate and diabetes prevalence had an inverse association with CFR. The association of CFR with the strictness of anti-COVID-19 measures was not found statistically significant, except for higher-income countries with active testing policies.

These and many other studies focused on analyzing the data that was available during the first months of the pandemic, also known as the COVID-19 “first wave”. At the end of 2020, several COVID-19 vaccines became available for the adult population. Massive vaccination campaigns were launched across the Globe in the hope of reaching “disease-free equilibrium”, where the majority of the population is immunized by a vaccine providing a long-term immunity with high efficacy, while providing “herd immunity” protection to those who cannot be immunized. Given the actual values of *R*_0_ and vaccine efficacy *VE*, one can calculate the herd immunity threshold of vaccinated individuals *f*_*v*_ using Eq. 2 (Gumel et al. 2021):

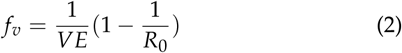

According to (Gumel et al. 2021) forecast, eliminating COVID-19 in the US using vaccination alone would require immunizing at least 70% of the US population by a vaccine of nearly 70% efficacy against infection. Otherwise, the epidemic will persist in the state of *endemic equilibrium*. The goal of our study is to explore the actual situation of COVID-19 dynamics up to September 30, 2021 in 50 countries having the highest absolute number of confirmed COVID-19 cases.

## Materials and methods

### Country-level data extraction

To explore the differences between countries, we have extracted the following 180-day moving averages of country-level factors from the Our World in Data COVID-19 dataset (Ritchie *et al*. 2020):

- Average of Delta: Average share of analyzed SARS-CoV-2 sequences that were the delta variant
- Average of total_cases_per_million: Average cumulative number of confirmed COVID-19 cases per one million people.
- Average of people_vaccinated_per_hundred: Average daily percentage of population vaccinated with any number of doses.
- Average of people_fully_vaccinated_per_hundred: Average daily percentage of fully vaccinated population.
- Average of total_boosters_per_hundred: Average daily percentage of population vaccinated with a booster dose.
- Average of stringency_index: The average daily value of the Government Response Stringency Index, a composite measure based on 9 response indicators including school closures, workplace closures, and travel bans.
- Average of population_density: Number of people divided by country’s area in square kilometers.
- Average of median_age: Median age of the country’s population.
- Average of aged_65_older: Share of the population that is 65 years and older.
- Average of gdp_per_capita: Gross domestic product at purchasing power parity.
- Average of cardiovasc_death_rate: Annual number of deaths from cardiovascular disease per 100,000 people.
- Average of diabetes_prevalence: Diabetes prevalence among people aged 20 to 79.
- Average of female_smokers: share of female smokers.
- Average of male_smokers: share of male smokers.
- Average of hospital_beds_per_thousand: Hospital beds per 1,000 people.
- Average of life_expectancy: Life expectancy at birth in 2019.
- Average of human_development_index: A composite index measuring three basic aspects of human development—a long and healthy life, knowledge and a reasonable standard of living.

### Estimating R and CFR

Our estimations of the average R and CFR values in each country are based on the daily values of confirmed COVID-19 cases and deaths reported by the Our World in Data website (Ritchie *et al*. 2020). The daily estimate of R on day *t* is calculated by Eq. 3.

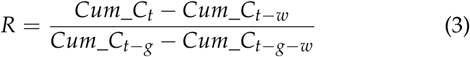

where *Cum*_*C*_*t*_ is the cumulative number of confirmed cases on day *t, w* = 7 days is the size of the sliding window, and *g* = 4 days stands for the average duration of the COVID-19 generation period (Last 2020).

The daily estimate of CFR on day *t* is calculated by Eq. 4.

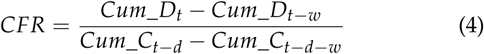

where *Cum*_*D*_*t*_ is the cumulative number of deaths on day *t* and *d* = 14 days represents the average time between testing positive and death (based on the COVID-19 Data Repository published by the Israeli Ministry of Health (of Health 2020)).

### Statistical analysis

For each country, we have calculated the daily moving averages of the reproduction number R using the pandas.DataFrame.rolling.mean() function with window = 180 days and min_periods (minimum number of observations in window required to have a value) = 30. The daily values of R were estimated from the first date when the total number of confirmed COVID-19 cases in the country has reached 1,000 until September 30, 2021. The daily moving averages of the case fatality rate CFR were calculated using Eq. 4, where the sliding window *w* was taken as the minimum between 180 days and the number of days since the date when the total number of COVID-19-related deaths in the country has reached 100. All missing values of country-level variables were imputed by the averages of the known values in the corresponding columns using the pandas.DataFrame.fillna function with default settings.

Pearson product-moment correlation coefficients between country-level variables were calculated using the scipy.stats.pearsonr function. Highly correlated variable pairs (Pearson’s *r* ≥ 0.60, *p* < 0.001) were excluded from further regression analysis. For the remaining variables, we have applied the forward-backward feature selection procedure based on p-value from the statsmodels.api.OLS function, which builds a multivariate linear regression model using the ordinary least squares (OLS) method. This stepwise variable selection procedure has two thresholds: threshold_in < threshold_out. It starts with only the intercept and at each step, it adds the most significant variable to the model. The selected variable should have the lowest p-value, which does not exceed threshold_in. The procedure stops when no variable meets the threshold_in criterion. In addition, at each step, the algorithm re-calculates the p-values of all existing model terms and removes the variables if their p-values exceed threshold_out. In our analysis, we set threshold_in to 0.05 and threshold_out to 0.10.

## Results

### Descriptive statistics

The descriptive statistics of all data variables is shown in Fig. 1. Fig. 2 shows Pearson’s correlation coefficients of all pairs of potentially predictive factors. No pairs of highly correlated variables (Pearson’s *r* ≥ 0.60, *p* < 0.001) were retained for further analysis. Thus, we have removed the following factors:

**Figure 1.**
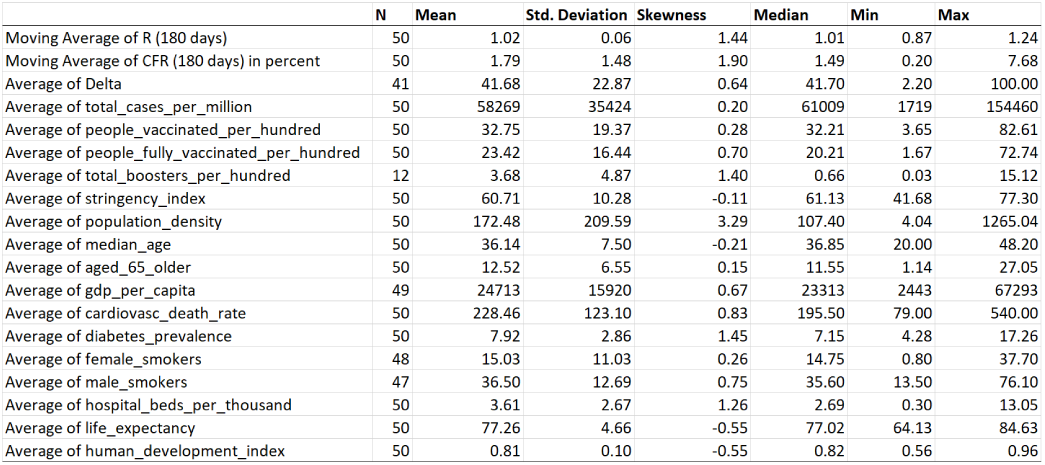
Descriptive statistics.

**Figure 2.**
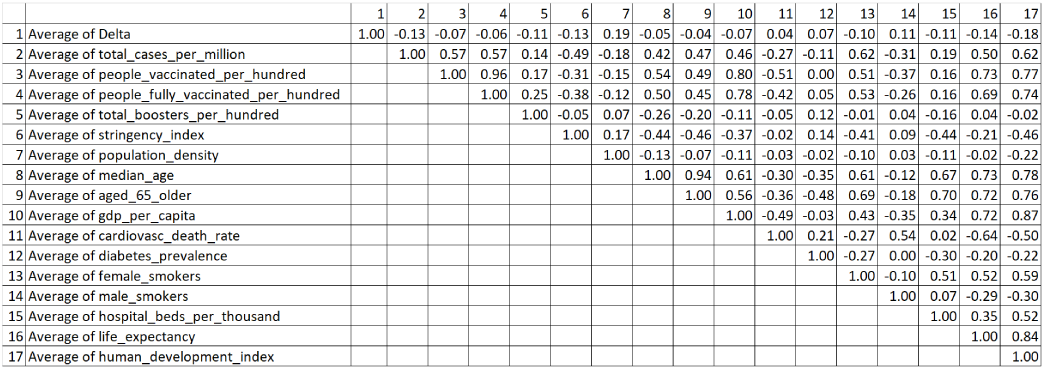
Pearson’s correlation coefficient.

- Average of people_vaccinated_per_hundred (highly correlated with Average of people_fully_vaccinated_per_hundred)
- Average of aged_65_older, Average of gdp_per_capita, Average of female_smokers, Average of hospital_beds_per_thousand, Average of life_expectancy, and Average of human_development_index (highly correlated with Average of median_age)

### R and CFR evolution over time

Figures 3 and 4 show the evolution of the 180-day moving averages for the country-level values of R and CFR, respectively. While both these parameters were highly unstable during the first months of the pandemic, their average values declined to much lower levels around July - October 2020 and remained stable since then. The differences between various countries, in terms of both parameters, have decreased over time as well. The mean values of country–level moving averages of R and CFR went down from 1.118 and 6.3%, respectively, on June 30, 2020 to 1.083 and 3.6% on September 30, 2020 and to 1.015 and 1.8% by September 30, 2021. In parallel, the 10% to 90% interpercentile range of R and CFR moving averages decreased from 0.288 and 13.3%, respectively, on June 30, 2020, to 0.151 and 7.7% on September 30, 2020, and to 0.107 and 3.3% by September 30, 2021.

**Figure 3.**
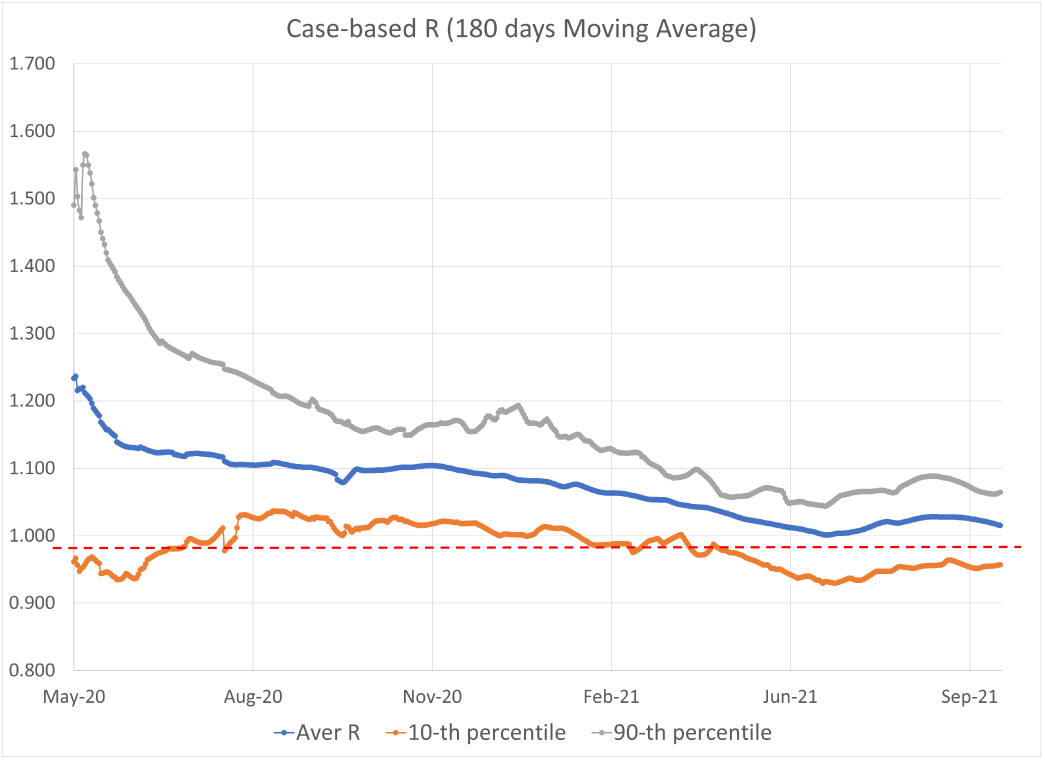
Moving Average of R (180 days).

**Figure 4.**
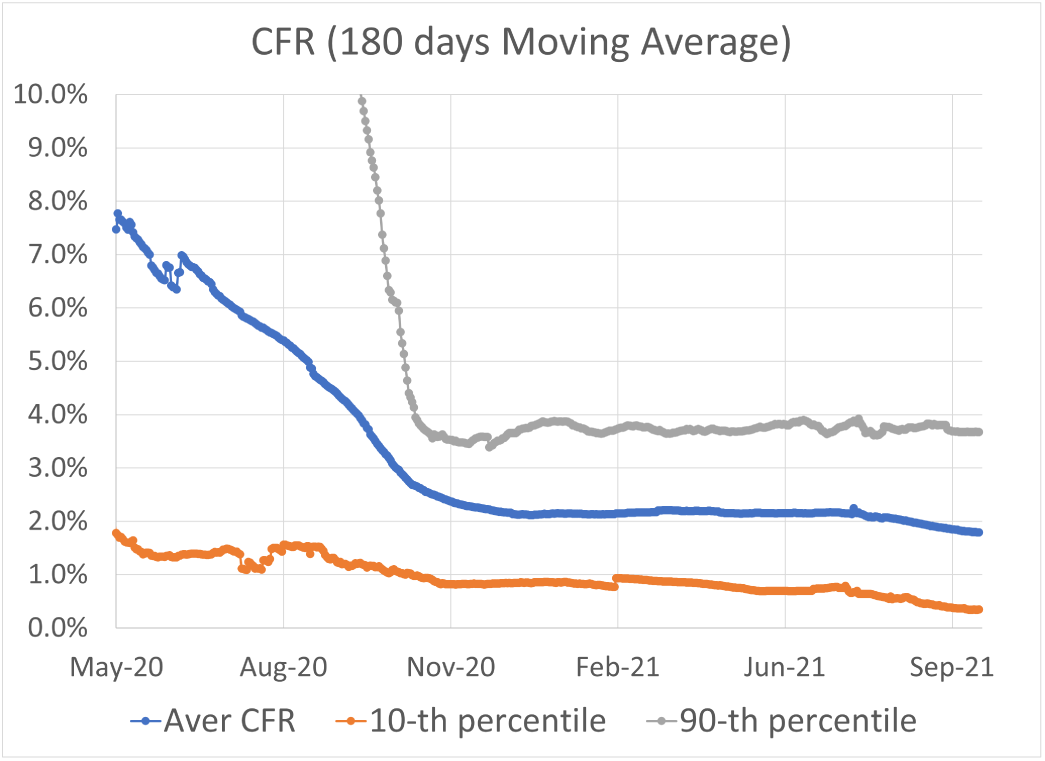
Moving Average of CFR (180 days).

### Country-level factors associated with R

After removing highly correlated variables, we have applied the forward-backward feature selection procedure with thresh-old_in = 0.05 and threshold_out = 0.10 in order to find a minimal set of factors significantly associated with the 180-day moving average of the reproduction number R calculated for each country on September 30, 2021. In the first feature selection step, the following variables had the lowest p-values (below 0.10):

- Average of Delta (p-value = 0.006, slope = 0.0010)
- Average of total_cases_per_million (p-value = 0.010, slope = -5.73e-07)

Consequently, Average of Delta was selected as the first variable to be added to the regression model. According to this model, an increase of 1% in the Delta variant share is associated with an increase of 0.001 (95% CI 0.000 to 0.002) in the average Reproduction Number R. A model based on the second significant factor (Average of total_cases_per_million) indicates that an increase of 1% in the total percentage of confirmed COVID-19 cases per country’s population is associated with a decrease of 0.006 (95% CI 0.001 to 0.011) in the average R. The effect of the Delta variant and confirmed cases on R is shown in Figures 5 and 6, respectively.

**Figure 5.**
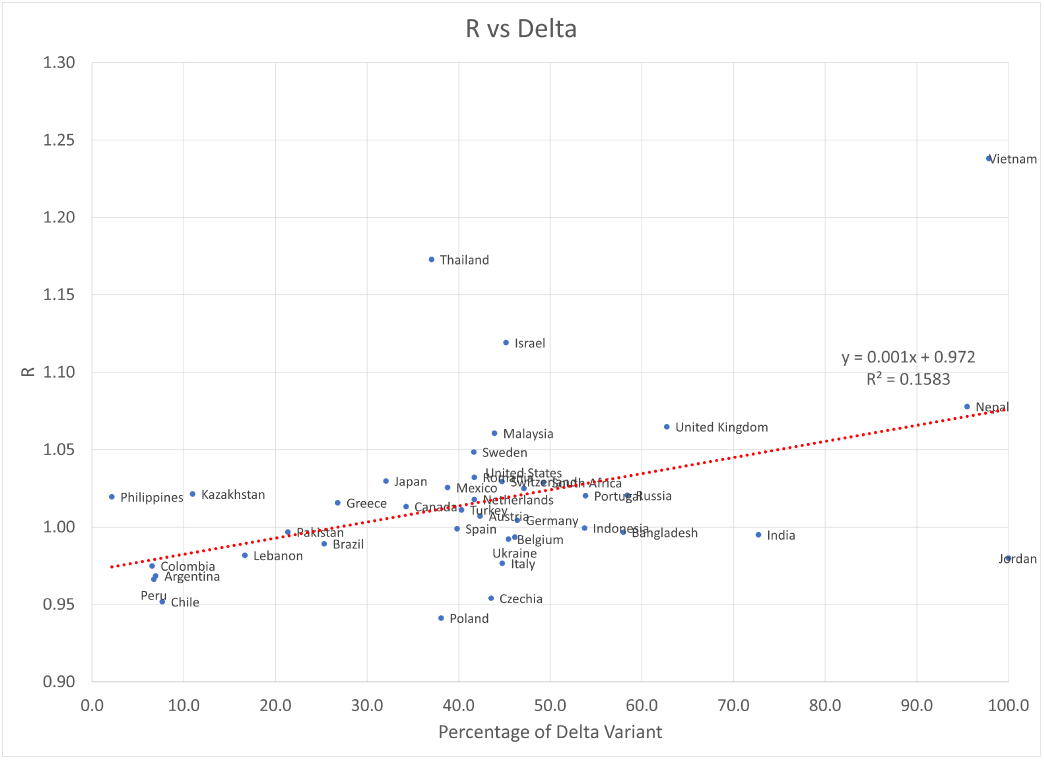
The effect of Delta variant on R.

**Figure 6.**
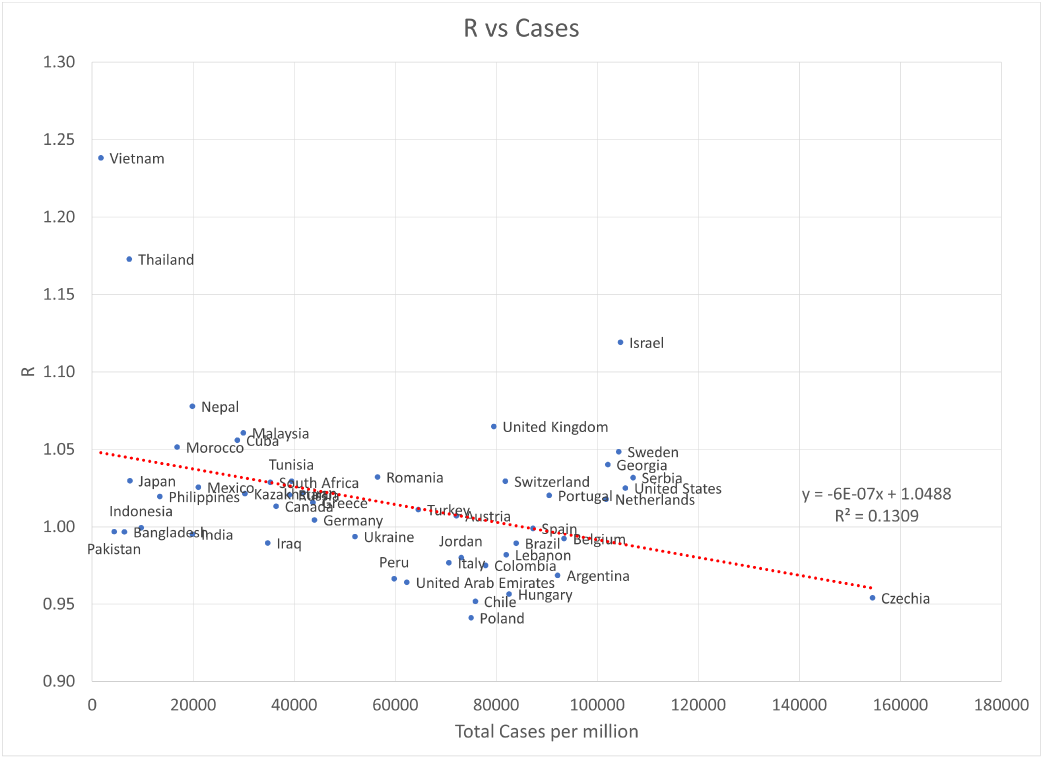
The effect of total confirmed cases on R.

In the second step, only Average of total_cases_per_million had p-value below 0.05 (0.02 < 0.05, slope = -5.018e-07) and thus it was selected as the second regression variable. No further variables were found statistically significant in the third step. The complete output of the resulting regression model is shown in Figure 7.

**Figure 7.**
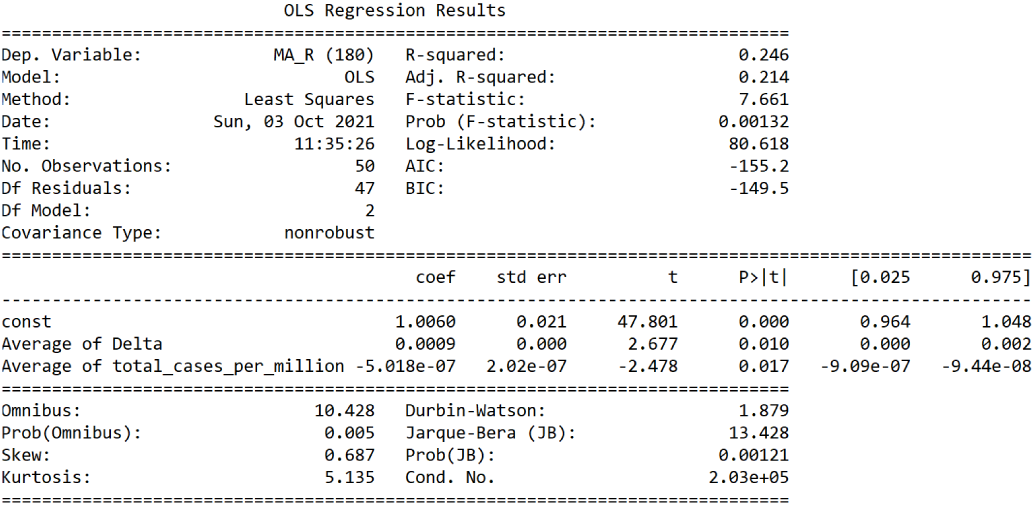
OLS Regression Results - R.

### Country-level factors associated with CFR

We have also applied the forward-backward feature selection procedure with threshold_in = 0.05 and threshold_out = 0.10 to find a minimal set of factors significantly associated with the 180-day moving average of the case fatality rate CFR calculated for each country on September 30, 2021. In the first step, only one variable, Average of people_fully_vaccinated_per_hundred, was found statistically significant (p-value = 0.003, slope = -0.036) and added to the regression model. According to this model, an increase of 1% in the total percentage of fully vaccinated people per country’s population is associated with a decrease of 0.04% (95% CI 0.01% to 0.06%) in the average CFR. The effect of vaccination percentage on CFR is shown in Figure 8.

**Figure 8.**
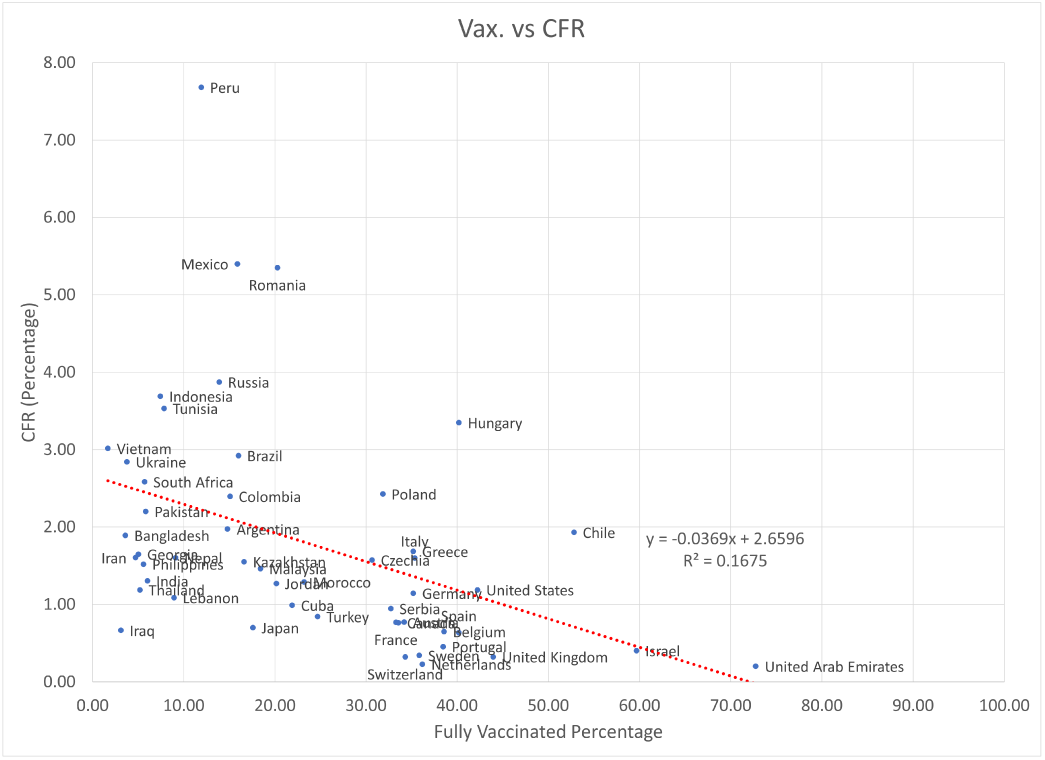
The effect of vaccination percentage on CFR.

In the second step, no further variables were were found statistically significant. The complete output of the resulting regression model is shown in Figure 9.

**Figure 9.**
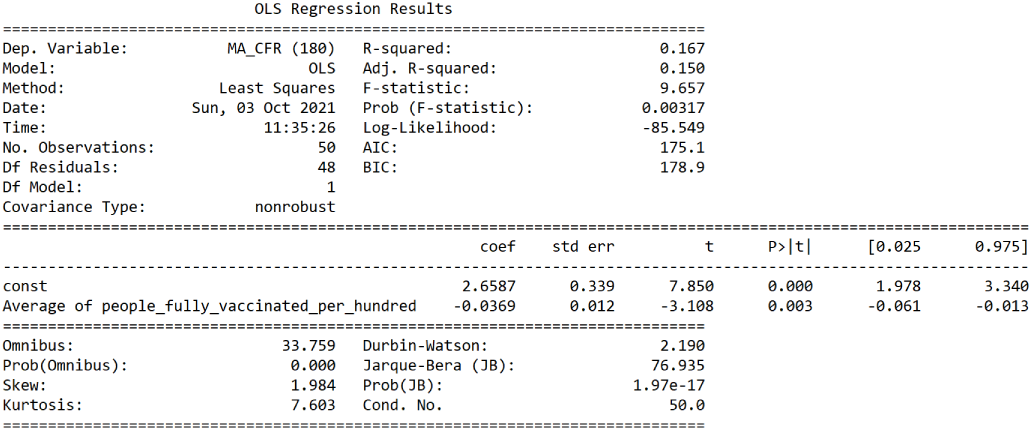
OLS Regression Results - CFR.

## Discussion

### Global COVID-19 equilibrium

The increasingly low levels of the cross–country variance of R and CFR (see Figures 3 and 4), along with the average value of R approaching the value of 1.0, may indicate that the COVID-19 pandemic has reached its point of a stable endemic equilibrium (Ashby and Best 2021). According to the mathematical model of COVID-19 presented in (Ahmed *et al*. 2021), a stable endemic equilibrium is maintained as long as the basic reproduction number *R*_0_ is greater than 1. In contrast, a stable disease-free equilibrium is achieved only when *R*_0_ < 1. Though we do not have a direct way of estimating the current value of *R*_0_ in each country, we may assume it to be close to the most recent peak in the effective reproduction number R. Considering the mean difference of 0.22 between the average and the maximum country-level values of R during our period of interest (April - September 2021), we should be able to reach a stable disease-free equilibrium of COVID-19 only after the average R will go *globally* below 1.0 − 0.22 = 0.78.

What is the required percentage of infected andor vaccinated population to make such an equilibrium possible? According to a regression model based on the Delta variant share and the number of confirmed cases (see Figure 7), *R* = 1.006 + 0.0009*Averageo f Delta −* 5.018*E −* 07*Averageo f total*_*cases*_*per*_*million*. Assuming the maximum share of the Delta variant (100%), the requirement of *R* = 0.78 implies *Averageo f total*_*cases*_*per*_*million* = (1.006 + 0.0009* 100 − 0.78)/5.018*E −* 07 = 635000*permillion* = 63.5%, which is much higher than any country’s current exposure to the virus of less than 22% and thus cannot be achieved globally in the foreseeable future. Moreover, reaching this level of natural herd immunity would require a prohibitively high cost in terms of human life. Thus, unfortunately, the regression model based on the data available at the end of September 2021 suggests that the endemic equilibrium of COVID-19 will maintain its stability as long as there will be no “game changer” in the form of either a vaccine more effective against infection or a less infectious mutant.

We may also estimate the average percentage of fully vaccinated population, which should bring the CFR close to the level of a seasonal flu (about 0.1%). According to the regression model shown in Figure 9, *CFR* = 2.6587 − 0.0369*Averageo f people*_ *f ully*_*vaccinated*_*per*_*hundred*. Setting CFR to 0.1 implies *Averageo f people*_ *f ully*_*vaccinated*_*per*_*hundred* = (2.6587 − 0.1)/0.0369 = 69%, which is clearly a feasible number, already exceeded in several countries (Ritchie et al. 2020). Thus, the currently available vaccines can be effective in reducing the mortality from the existing COVID-19 variants close to the level of a seasonal flu.

### Demographics and COVID-19

Our regression analysis has shown that the median age and the average density of a country’s population are not statistically significantly associated with the average reproduction number. Contrary to (Ahammed et al. 2021), we have also found no statistically significant associations of any demographic parameter with CFR.

### Economic development and stringency measures

Similar to the findings of (Cao *et al*. 2020), the association of the Average of stringency_index with R and CFR was not found statistically significant. Thus, we could not identify any statistically significant effects of government measures restricting the people’s behavior (such as lockdowns) on COVID-19 dynamics. The economic development factors (gdp_per_capita, hospital_beds_per_thousand, life_expectancy, and human_development_index) were removed from the regression analysis due to their high correlation with the median age.

### Strengths and limitations

Contrary to numerous works analyzing the data accumulated during the first months of the pandemic, this study explores the long–term trends in country–level Reproduction Number and the Case Fatality Rate of COVID-19 from the beginning of the pandemic until September 2021. We also investigate the long–term statistical dependence of the COVID-19 Reproduction Number and the Case Fatality Rate on epidemiological, demographic, economic, immunization, and government policy factors in each country. The findings of this study may have important implications for the health authorities worldwide in considering their vaccination policies, non-pharmaceutical intervention measures, and resource allocation decisions.

This retrospective study suffers from several limitations. First, the officially reported numbers of daily COVID-19 confirmed cases depend on the local testing policy and usually underestimate the true number of carriers in the population. As daily virus testing of the entire population is not possible, the actual reporting rate is usually unknown. Second, the officially reported numbers of daily COVID-19 deaths in some countries may include all deceased individuals who tested positive for COVID-19 (people who “died with coronavirus”), disregarding their actual cause of death, and exclude some victims (people who “died from coronavirus”), because they were not tested for COVID-19 before their death. The reported numbers depend on the time-changing policy of each country, which is not always made explicit to the public. Last but not least, the future dynamics of COVID-19 depends on the unknown characteristics of new variants, short and long term efficacy of currently developed vaccines, government decisions, public behavior, and other uncertainty factors. Drastic changes in some of these factors may render the models based on the past data completely useless.

## Conclusion

The continuous decrease in the country-level moving averages of R, down to the level of 1.0, accompanied by repeated outbreaks (“waves”) in various countries, may indicate that COVID-19 has reached its point of a stable endemic equilibrium. In our regression analysis, only the Delta variant share and the total percentage of confirmed cases were identified as statistically significant factors associated with the average values of R in different countries. According to the regression model shown in Figure 7, only a prohibitively high level of herd immunity (about 63%), associated with a tremendous cost in terms of human life, may naturally stop the endemic by reaching a stable disease-free equilibrium. On the other hand, the average percentage of fully vaccinated population, which appears to be statistically significantly associated with country-specific CFR, can bring it close to the level of a seasonal flu (about 0.1%) after vaccinating more than 70% of a country’s population. It is noteworthy that no statistically significant effect of vaccinations on R was found in our analysis. Thus, while the currently available vaccines prove to be effective in reducing mortality from the existing COVID-19 variants, they seem unlikely to stop the spread of the virus in the foreseeable future. In addition, the performed data analysis revealed no statistically significant effects of government measures restricting the people’s behavior (such as lockdowns) on either R, nor the CFR.

## Data Availability

All data used in this study is publicly available at https://ourworldindata.org/ and cited in the article.

## Patient consent for publication

Not required.

